# Adaptation and Validation of Brief Tablet-Based Cognitive Assessment Tool in Uganda

**DOI:** 10.64898/2026.09.21.26363152

**Authors:** Roslyn Valdespino, Gabrielle Hromas, Chen-Pin Wang, Robert Paul, Noeline Nakasujja, Zahra Reynolds, Flavia Atwiine, Edna Tindimwebwa, Meredith Greene, Eliza Passell, Christine S. Ritchie, Susanne S. Hoeppner, Alexander C. Tsai, Janet Seeley, Amy Werry, Emi Varfaj, Sudha Seshadri, Samson Okello, Stephen Asiimwe, Deanna Saylor, Katherine L. Possin, Elena Tsoy, Mark J. Siedner, Jeremy A. Tanner

## Abstract

**Background:** Sub-Saharan Africa (sSA) faces accelerated growth in Alzheimer’s disease and related dementias (ADRDs), including in older people living with HIV (PLWH). However, there are limited contextually relevant diagnostic tools to assess cognitive impairment in the region. The Tablet-based Cognitive Assessment Tool (TabCAT) is a digital platform hosting cognitive tests with automated scoring that can be administered by non-specialists and are designed for use across diverse cultures, languages, and education levels. We translated, adapted, and assessed the validity of a brief battery of TabCAT tests in the Uganda Aging and Dementia Cohort Study (UADCS), a prospective cohort of older PLWH and age- and sex-similar adults not living with HIV in southwestern Uganda.

**Methods:** Four TabCAT tests were translated and culturally adapted through expert review and focus groups of adults at study sites. Psychometric validity was examined by assessing floor and ceiling effects, association with known demographic predictors, and concurrent and divergent validity against a reference-standard cognitive testing battery previously validated and employed in Uganda. Z-scores on all tests were derived using a regression-based normative approach to adjust for age, sex, education, urbanicity, and literacy. Receiver Operating Characteristic (ROC) curves were fit to assess the TabCAT Composite Score performance in discriminating objective cognitive impairment (defined using Jak/Bondi criteria) in the total sample, then stratified by HIV serostatus.

**Results:** TabCAT tests were reported by local experts and focus groups to have acceptable face and content validity. Participants (n=563, mean age 60±6.4, 50% female, 51% did not complete primary school, 16% were not literate, 49% PLWH) completed the reference-standard and TabCAT battery. For TabCAT tests, there were no notable floor or ceiling effects, and test scores were associated with age and education as expected. Correlations between TabCAT and reference-standard tests were stronger in aligning cognitive domains (memory, executive function) than in non-aligning domains (motor). The TabCAT Composite Score discriminated cognitive impairment with good performance (c-statistic 0.77; 95% CI 0.72-0.81), including among PLWH.

**Discussion:** TabCAT tests demonstrated face, content, construct, concurrent, and criterion validity for measuring cognition and detecting objective cognitive impairment among older adults in Uganda, including PLWH. These results support the potential of tablet-based brief cognitive assessment tools to measure cognitive performance and detect cognitive impairment in Uganda, including among older PLWH, and in populations with lower levels of education and literacy.

## Introduction

Approximately 50 million people worldwide live with Alzheimer’s disease and related dementias (ADRDs), and this number is projected to triple in the coming decades.^1^ Sub-Saharan Africa (sSA) is anticipated to have the fastest growth in older adults globally, with most concentrated in rural regions.^1,2^ Concurrently, the life expectancy and number of older people living with HIV (PLWH) in sSA is also substantially increasing. This is in large part due to the success of prior public health initiatives, including increased access to HIV care, earlier diagnosis, and use of antiretroviral therapy (ART).^3–6^ HIV has been linked to cognitive impairment and dementia in up to 50% of PLWH, often presenting with clinical and cognitive testing that overlap with AD, though data on older well-treated PLWH in sSA is lacking.^7–9^ The intersection of aging, ADRDs, and HIV portends a uniquely pressing concern for cognitive decline in sSA.

Worldwide, an estimated 75% of people with ADRDs remain undiagnosed,^10^ well below the WHO Dementia Global Action Plan target of 50% diagnosis in at least 50 countries by 2025.^11^ Cognitive impairment and ADRDs remain severely underdiagnosed in the Global South, including in sSA. Comprehensive neuropsychological testing traditionally requires specialty training to administer and interpret results, and there are not enough specialists in sSA to meet the growing need.^12,13^ Most brief cognitive assessment tools, such as the Mini-Mental State Examination, were developed and validated in high-literacy populations and in the Global North, and perform poorly in lower education and culturally diverse populations, including in sSA.^12,14,15^ A lack of accessible tools to identify cognitive impairment in sSA limits efforts to advance ADRD-relevant research, care, interventions, and public health initiatives. There is an urgent public health need for scalable and cost-effective diagnostic tools to identify cognitive impairment that can be used globally, including in sSA and in older PLWH.

The Tablet-based Cognitive Assessment Tool (TabCAT) tests can be used by non-specialists to administer a brief, 10-minute cognitive assessment battery using a tablet with automated scoring provided.^16–19^ TabCAT tests are designed to minimize cultural bias across diverse global populations, including individuals with lower levels of formal education. Prior studies have demonstrated that brief TabCAT batteries can accurately detect cognitive impairment, discriminate amyloid positive from negative individuals, and generalize to low-education populations and those with no prior tablet experience.^16–19^ A study in Cuba demonstrated that TabCAT tests had better performance than the Montreal Cognitive Assessment (MoCA) in identifying clinically diagnosed mild cognitive impairment after translation and cultural adaptation.^18^ However, TabCAT has not previously been validated against reference-standard cognitive testing in populations in sSA or in older PLWH, where such brief cognitive assessment tools are needed to support diagnosis and care.

Our overall objective was to translate, culturally adapt, and assess the validity of TabCAT as a digital cognitive assessment tool to detect cognitive impairment in a representative cohort in rural Uganda, including older PLWH. Specifically, we aimed to (1) translate and culturally adapt the TabCAT test battery; (2) validate the performance of the TabCAT subtests against reference-standard, pen-and-paper neuropsychological tests; and (3) assess the discriminative ability of the TabCAT to detect objective cognitive impairment, including in PLWH. We sought to assess TabCAT as a promising brief digital cognitive assessment tool for further use in populations in sSA and in PLWH.

## Methods

### Study Design and Cohort

This was a cross-sectional analysis of data collected from participants in the Uganda Aging and Dementia Cohort Study (UADCS) during the third annual (2023) study visit. The UADCS is a prospective cohort study of older adults in southwestern Uganda designed to identify determinants of quality of life, aging, and cognitive functioning in older adults in Uganda, with a focus on the contributions of chronic, well-treated HIV.^20–22^ The cohort includes equal proportions of PLWH and older adults without HIV and of men and women. The cohort initially enrolled 297 PLWH on ART from HIV clinics, and 302 older adults not living with HIV from the community using population-based sampling approaches. PLWH had to be >48 years old, on ART for a minimum of three years, and reside within a 20-km radius of HIV clinic sites. Age- and sex-similar older adults not living with HIV were selected from the clinic catchment areas using population census data and were enrolled in a 1:1 ratio. Study visits took place at two sites in southwestern Uganda: the semi-urban Mbarara University of Science and Technology (MUST) in Mbarara and the rural Kabwohe Clinical Research Centre (KCRC) in Kabwohe. Participants underwent annual assessments of lifestyle, cardiovascular health, inflammatory markers, cognitive function, and physical functional measures.^23^

### Reference-Standard Neuropsychological Testing (Pen-and-Paper) Administration

UADCS participants complete the Ugandan Neuropsychological Battery as part of the annual study visits beginning in the second year of the cohort. The Uganda Neuropsychological Battery was originally developed to assess HIV-associated cognitive impairment. The battery consists of measures of learning and memory (WHO/UCLA AVLT^24^), attention (Color Trails Part 1;^25^ Digit Span Forward^26^), executive function (Color Trails Part 2;^25^ Symbol Digit Modalities Test (written form);^27^ Digit Span Backwards^26^), language (Verbal Fluency – Animals^28,29^), fine motor speed (Finger Tapping;^30,31^ Grooved Pegboard^32^), and gross motor speed (Timed Gait).^33,34^ This battery was previously translated into the local-language Runyankore and successfully implemented in prior studies of PLWH in Uganda.^35–38^ The tests were administered by research staff after undergoing training and certification with local and international neuropsychologists with routine monitoring.

### TabCAT Translation, Adaptation, and Administration

The TabCAT testing battery consisted of 4 tests: Favorites (multimodal associative memory task where individuals are asked to learn and remember the association between a face and favorite food and animal), Birdwatch (visual associative memory task where individuals are asked to learn and remember the association between a bird and a landscape), Flanker (executive function and inhibition task where individuals are asked to indicate the direction of the arrow in the center among congruent or incongruent surrounding arrows), and Match (executive function and processing speed test where individuals are asked to match a number with a symbol as quickly as possible). For Flanker, participants were required to pass a training trial for data to be recorded.

For translation, the TabCAT test battery was translated into Runyankore and then back translated to English by independent expert interpreters. Next, a group of 5 professionals with expertise in cognitive testing and fluency in both Runyankore and English reviewed and finalized test instructions and item content to optimize functional equivalence and cultural appropriateness of the translated versions of the tests. We adapted previously described procedures to identify which animals and foods to include in Favorites.^18,19^ In brief, 5 native speakers ranked the 20 most common animals and the 20 most common fruits and vegetables in the language. We then averaged the word frequency ranking for each word and excluded (a) the 2 most frequent words from each category; (b) long, complex, or compound words, and (c) words only listed by 1 participant. We then created 4 alternate forms for animal and food word stimuli matched on word frequency and syllable count.

The translated TabCAT test battery was then administered to 5-7 local volunteers across age at each study site by trained research staff. Following administration, focus groups were conducted at each study site to assess cultural appropriateness, clarity, design, difficulty, length, and usability of the TabCAT tests. Each focus group comprised of 5–7 middle-aged and older adults balanced by gender and representing a range of educational backgrounds. The focus groups were conducted by a trained bilingual facilitator with a note-taker and observer present. The TabCAT tests were further adapted based on participant feedback, and re-training of research staff was performed. An initial pilot study was conducted in 50 participants for quality control. TabCAT was then incorporated into the cognitive testing in annual UADCS visits starting in Year 3 and administered alongside the Ugandan Neuropsychological Battery (same day).

### Covariates

HIV infection status was confirmed via annual HIV testing among those without HIV. Demographic variables used to develop regression-based norms included age (self-reported and confirmed on government identification, continuous variable), sex (self-reported, male/female), education (self-reported highest level of educational attainment, categorical variable including no schooling, some primary school, completed primary school, some secondary school, and completed secondary school or higher), urbanicity (defined based on study site using the Uganda Bureau of Statistics,^39^ rural/semi-urban), and literacy (yes/no). Literacy was defined as being able to read and write and determined by self-report and then confirmed by research staff during cognitive testing. Literacy in the cohort had high concordance with separate questions on whether the participant was able to read from 1 to 25 (97% agreement) and count from 1 to 25 (90% agreement). As a result, Color Trails Part 1 and 2, Symbol Digit Modalities, and TabCAT Match were not included in analyses in participants who were not literate, as these tests involved reading numbers and/or letters.

### Regression-Based Normative Z-scores

To account for sociodemographic influences on cognitive test performance, we generated regression-based normative Z-scores for both the reference-standard and TabCAT cognitive tests in the third annual study visit using a cognitively unimpaired subset of participants. Because no external normative data were available for older adults in Uganda, we derived an internal reference sample of individuals not living with HIV who reported no history of conditions that could potentially affect cognitive performance (e.g., prior tuberculosis, stroke, severe traumatic brain injury (TBI; defined as TBI with loss of consciousness >24 hours using the Ohio State University TBI Identification Method^40^), or any subjective cognitive concerns (defined as a score of ≥8 on the World Health Organization Disability Assessment Schedule WHODAS 2.0 cognition subscore^41^). A multidisciplinary expert review panel consisting of a neuropsychologist, cognitive neurologist, and biostatistician reviewed available records and examiner reports of remaining participants to exclude individuals with reported concerns for cognitive impairment. Of the initial cohort (n=563), 231 participants were included in the final normative sample as cognitively unimpaired (**Supplementary Table 2**).

Within this normative sample, we evaluated potential covariates associated with cognitive performance. Age, sex, education, literacy, and urbanicity (site) were consistently and independently associated with performance across both test batteries and were included in regression models. There were significant interactions between participant sex and other covariates, therefore sex-stratified equations were used for modeling. Depression, anxiety, and alcohol use were explored but not meaningfully associated with test performance and were not included in the final regression model.

Regression-based Z-scores were then calculated for all participants in the UADCS using Formulas A and B after stratifying by sex (due to significant interactions between sex and other covariates). Among binary covariates, the reference group for urbanicity was the semi-urban site (Mbarara) and for literacy was non-literate. Education was treated as ordinal. Z-scores were coded such that higher scores reflected better cognitive performance, and lower scores reflected weaker performance.

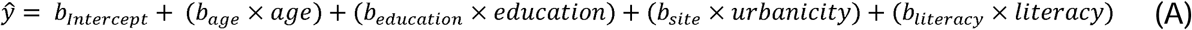

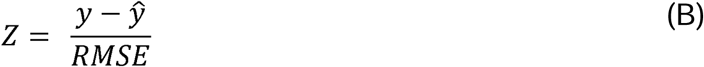

Additional details are available in the Supplementary Materials.

### TabCAT Predictors

TabCAT test scores were used as continuous predictors. For each participant, regression-based Z-scores were calculated for each TabCAT test: Favorites (total correct across all trials; min=0, max=24), Birdwatch (total correct; min=0, max=56), Flanker (total score; min=0, max=10), and Match (total correct; min=0, no max (continuous)). The TabCAT Composite Score was defined as an individual’s mean Z-score across all available TabCAT subtests.

### Reference-Standard Outcomes

Similarly, Z-scores were calculated for each test in the Ugandan Neuropsychological Battery test and analyzed as continuous outcomes. Individual test scores included the WHO/UCLA AVLT trials 1-5 total score and Delayed Recall, Color Trails Test Parts 1 and 2 (seconds), Symbol Digit Modalities Test total score, Forward and Backward Digit Span, Animal Verbal Fluency (total correct), Finger Tapping (dominant hand, mean of 5 trials), Grooved Pegboard (dominant and non-dominant hand, seconds), and Timed Gait (seconds, mean of 3 trials). A Global Cognitive Composite Score was calculated for each participant as the mean Z-score across tests spanning learning and memory, attention, executive function, and language.

The primary binary outcome was objective cognitive impairment defined using Jak/Bondi criteria, which has previously been validated in both clinic and population-based studies.^42–44^ Objective cognitive impairment was defined as ≥2 test scores at least 1 standard deviation below the mean in ≥1 cognitive domain, or ≥1 test score at least 1 standard deviation below the mean in ≥3 domains. Cognitive domains included learning and memory, attention, executive function, and language. Fine and gross motor domains were not included.

In sensitivity analyses, objective cognitive impairment was defined per Jak/Bondi criteria with a more stringent threshold of -1.5 standard deviations. Additional definitions used the Global Cognitive Composite Score with 4 separate cut points of increasing severity: -0.5, -0.75, -1.0, and -1.5.

### Statistical analysis

#### Psychometric validity

To assess construct validity, we first evaluated the distribution of raw TabCAT test scores to assess floor and ceiling effects. We then performed unadjusted linear regression to separately assess the association of TabCAT test scores with age and education. Concurrent validity was assessed using Spearman rank correlations between TabCAT tests and reference-standard tests in aligning cognitive domains (e.g., Favorites and WHO/AVLT Total Learning and Delayed Recall). Divergent validity was assessed by Spearman correlations between TabCAT tests and reference-standard tests in non-aligning domains (e.g., Favorites and Timed Gait). Primary analyses used

Z-scores, and sensitivity analyses used raw test scores.

#### Discrimination of Cognitive Impairment

We evaluated the performance of the TabCAT Composite Score to discriminate objective cognitive impairment (per Jak/Bondi criteria on reference-standard tests) using receiver operating characteristic curve analyses and the area under the curve (AUC) in the total cohort. We then compared the performance in PLWH compared to HIV negative participants using Delong’s test. In sensitivity analyses, we assessed the discriminative performance of TabCAT Composite Score using alternative definitions of objective cognitive impairment. Additionally, we evaluated AUCs for individual TabCAT subtests.

Analyses were performed using SAS software, version 9.4 (SAS Institute Inc., Cary, NC, USA), and data visualization was performed using the ggplot2 package in R.^45^

### Ethical Approval

Ethical approval was obtained from Massachusetts General Hospital Institutional Review Board (IRB), the Mbarara University of Science and Technology Research Ethics Committee (MUSTREC), and the Uganda National Council for Science and Technology (UNCST).

## Results

### Participants

At study year 3, there was >93% retention of the initial cohort (n=563). Characteristics of participants in the total cohort are detailed in **Table 1** and **Supplementary Table 1**, and of the normative sample in **Supplementary Table 2**. A total of 563 participants completed annual visits including cognitive testing in study year 3 at two study sites in southwestern Uganda. The mean age was 60.4 years (range 50-90), 50% were female, and 49% were PLWH. Most participants had less than a primary school education (51%), and 16% (n=89) were unable to read and write. Most participants were cognitively unimpaired (n=445, 79%) per Jak/Bondi criteria using the Ugandan Neuropsychological Battery.

**Table 1.** Participant Characteristics in the Uganda Aging and Dementia Cohort Study.

| Characteristic | Total Cohort<br>(n=563) |
| --- | --- |
| Age in years, mean (SD; range) | 60.4 (6.4; 50 to 90) |
| Female, n (%) | 284 (50%) |
| Education, n (%) |  |
| No schooling | 83 (15%) |
| Some primary school | 203 (36%) |
| Completed primary education | 130 (23%) |
| Some secondary school | 81 (14%) |
| Secondary school or higher | 66 (12%) |
| Literacy, n (%) |  |
| No | 89 (16%) |
| Yes | 474 (84%) |
| Urbanicity, n (%) |  |
| Semi-urban (Mbarara) | 233 (41%) |
| Rural (Kabwohe) | 330 (59%) |
| HIV Positive, n (%) | 274 (49%) |
| Global Cognitive Composite, Z-score mean (SD; range) | -0.03 (0.7; -2.6 to 1.9) |
| Objective cognitive impairment<sup>1</sup>, n (%) | 118 (21%) |
| TabCAT Cognitive Composite, Z-score mean (SD; range) | -0.05 (0.7; -1.9 to 2.4) |
| Favorites, Z-score mean (SD; range) | -0.05 (1.0; -2.5 to 3.0) |
| Birdwatch, Z-score mean (SD; range) | -0.03 (1.0; -2.4 to 5.2) |
| Flanker, Z-score mean (SD; range)<sup>2</sup> | -0.1 (1.1; -4.4 to 2.6) |
| Match, Z-score mean (SD; range)<sup>3</sup> | -0.02 (1.0; -2.9 to 3.1) |

1 = Defined per Jak/Bondi criteria
2 = n=461 as participants unable to complete the training trial were excluded
3 = n=474 as participants who were unable to read/write were excluded

### TabCAT Tests

TabCAT Flanker results were available in 461 participants who completed the training trial, and TabCAT Match in 474 participants with literacy. The final regression models for calculating Z-scores for each TabCAT test are displayed in **Supplementary Table 3**. The TabCAT Composite Score was derived from the mean Z-score of 4 completed TabCAT tests in 414 participants (74%), from 3 completed tests in 107 (19%) participants, and from 2 completed tests in 42 (7%) participants.

### Face and Content Validity

Overall, TabCAT tests demonstrated acceptable face and content validity per local bilingual translators and focus groups at both sites. Focus group participants generally found TabCAT assessments appropriate and understandable, with most reporting the instructions were clear and some expressing positive engagement (e.g., “this is a very good tool to open up the mind”). The most consistent feedback was a request for additional time to complete tasks and explicit instructions and reminders when tests emphasize speed in addition to accuracy. Minor revisions to instructions and translations were suggested to improve clarity. Among TabCAT tests, Match and Flanker were consistently described as clear and appropriate, while Favorites raised concerns regarding cultural representativeness of the faces locally.

### Psychometric Validity

We evaluated construct validity by assessing the association between TabCAT tests and known demographic predictors. Increasing age was associated with lower raw scores on each TabCAT test (**Figure 1**). Higher levels of educational attainment were associated with higher raw scores on each TabCAT test (**Figure 2**). Relationships between TabCAT test scores and additional demographic variables used in deriving Z-scores, including sex, urbanicity, and literacy, are shown in **Supplementary Figures 1–3**.

**Figure 1.**
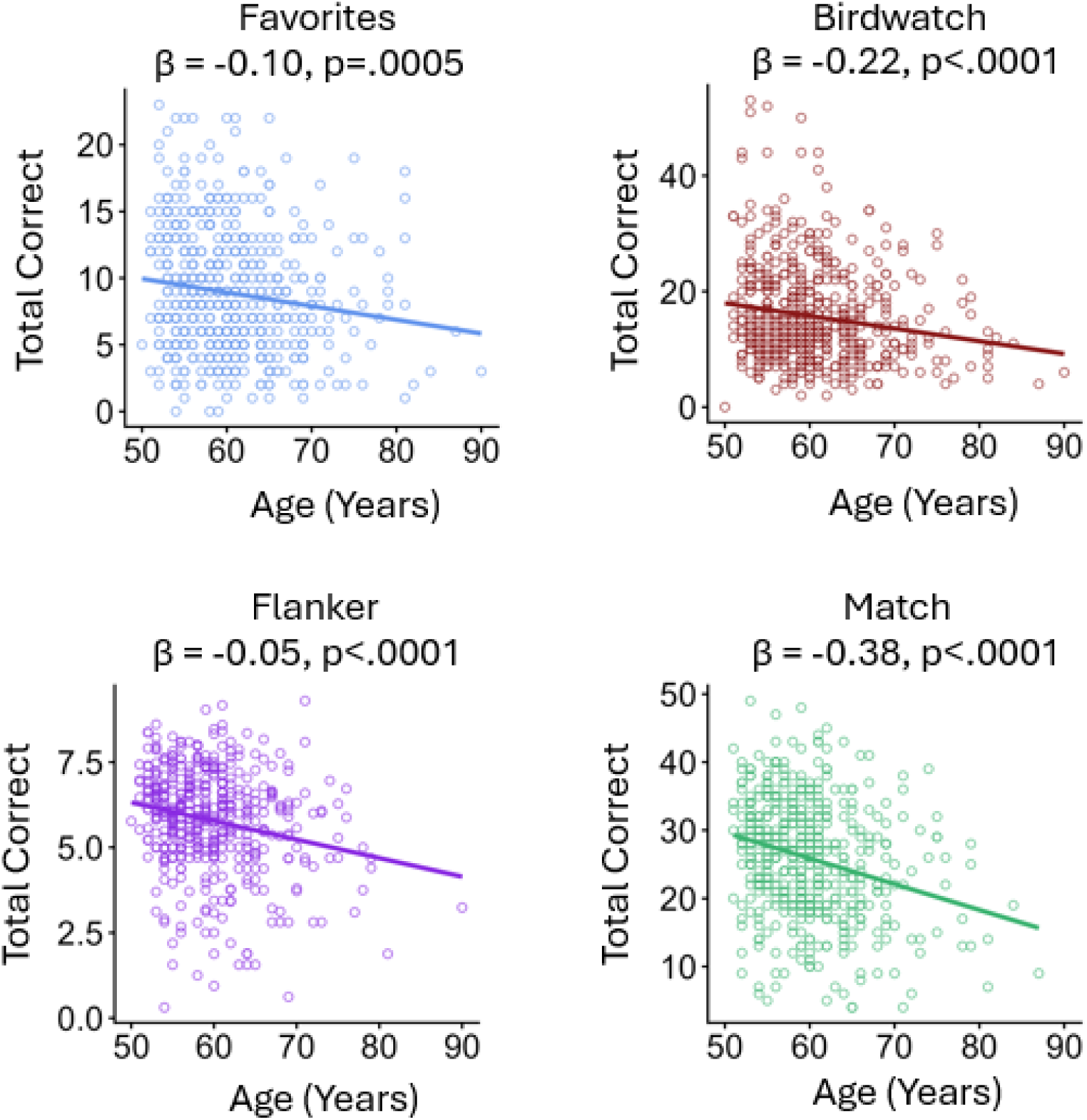
Age association with TabCAT test performance. Increasing age was associated with a decline in raw scores for each TabCAT test.

**Figure 2.**
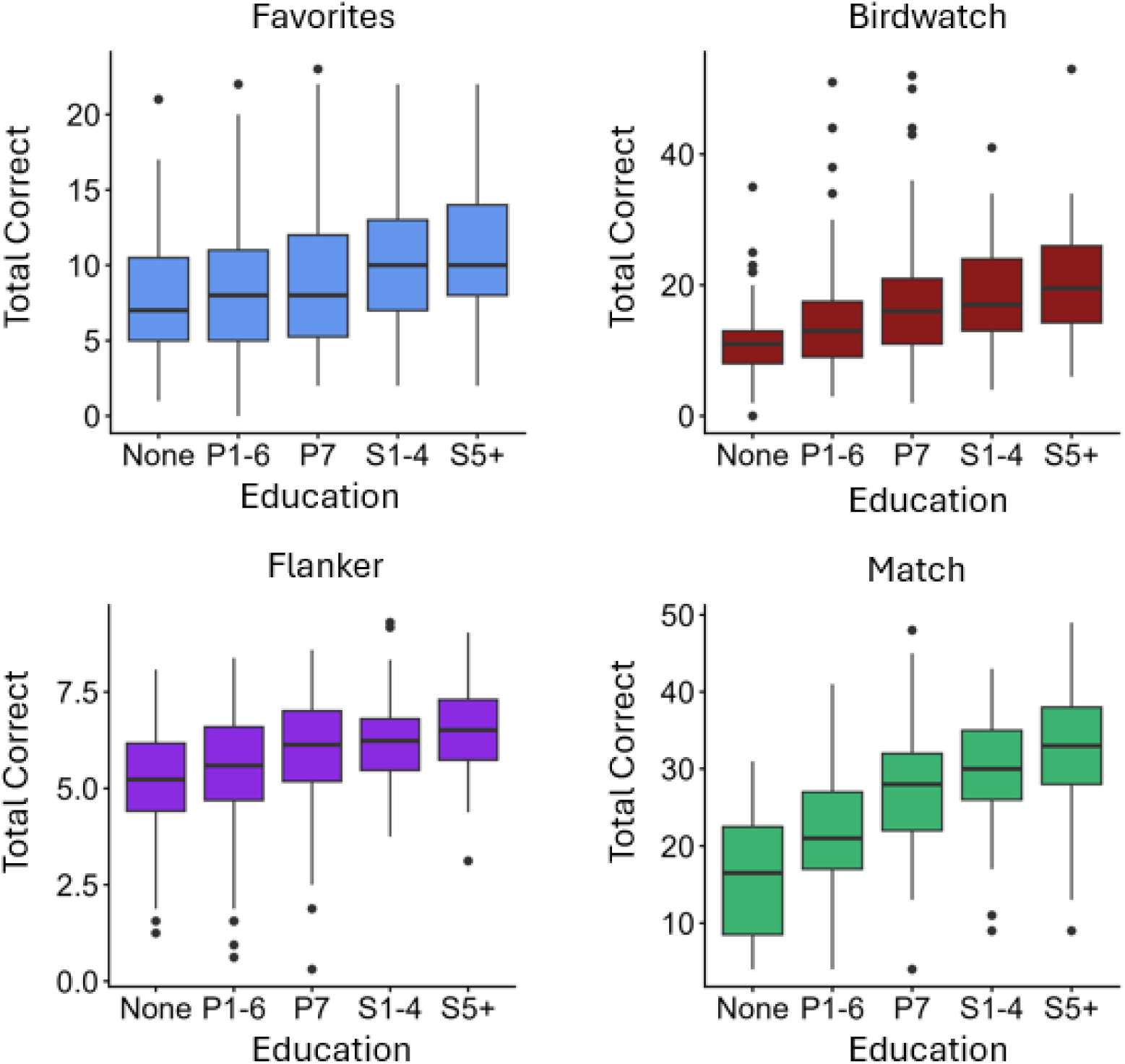
Education association with TabCAT Test Performance. Higher levels of educational attainment are associated with higher raw scores in each TabCAT subtest. Abbreviations: P1-6 = 1 to 6 years of primary school; P7 = completed primary school; S1-4 = 1 to 4 years of secondary school; S5+ completed secondary school or greater

There did not appear to be notable floor or ceiling effects for any TabCAT test in the cohort. For each test, fewer than 2% of participants achieved a score at the minimum or maximum possible value, or within one point of those values (e.g., scores of 23–24 on Favorites, where 24 is the maximum score). These findings were consistent after stratifying by literacy (**Supplementary Table 4**).

Correlations between TabCAT and reference-standard tests in aligning and non-aligning domains are shown in **Figure 3**. Favorites, Birdwatch, and Flanker demonstrated significant but weak correlations with reference-standard tests in aligning domains, whereas Match showed strong correlations with executive function measures. Correlations with non-aligning (motor) domains appeared weaker than correlations with aligning domains for each TabCAT test, though many remained statistically significant. Match had the strongest correlations with corresponding measures (Symbol Digit Modalities: ρ=0.63, p<.001; Color Trails Part 2: ρ=0.58, p<.001), while Favorites showed the weakest correlations with reference-standard memory measures (Delayed Recall: ρ=0.15, p<0.001; Total Learning: ρ=0.19, p<.001). Results were similar in sensitivity analyses using raw test scores.

**Figure 3.**
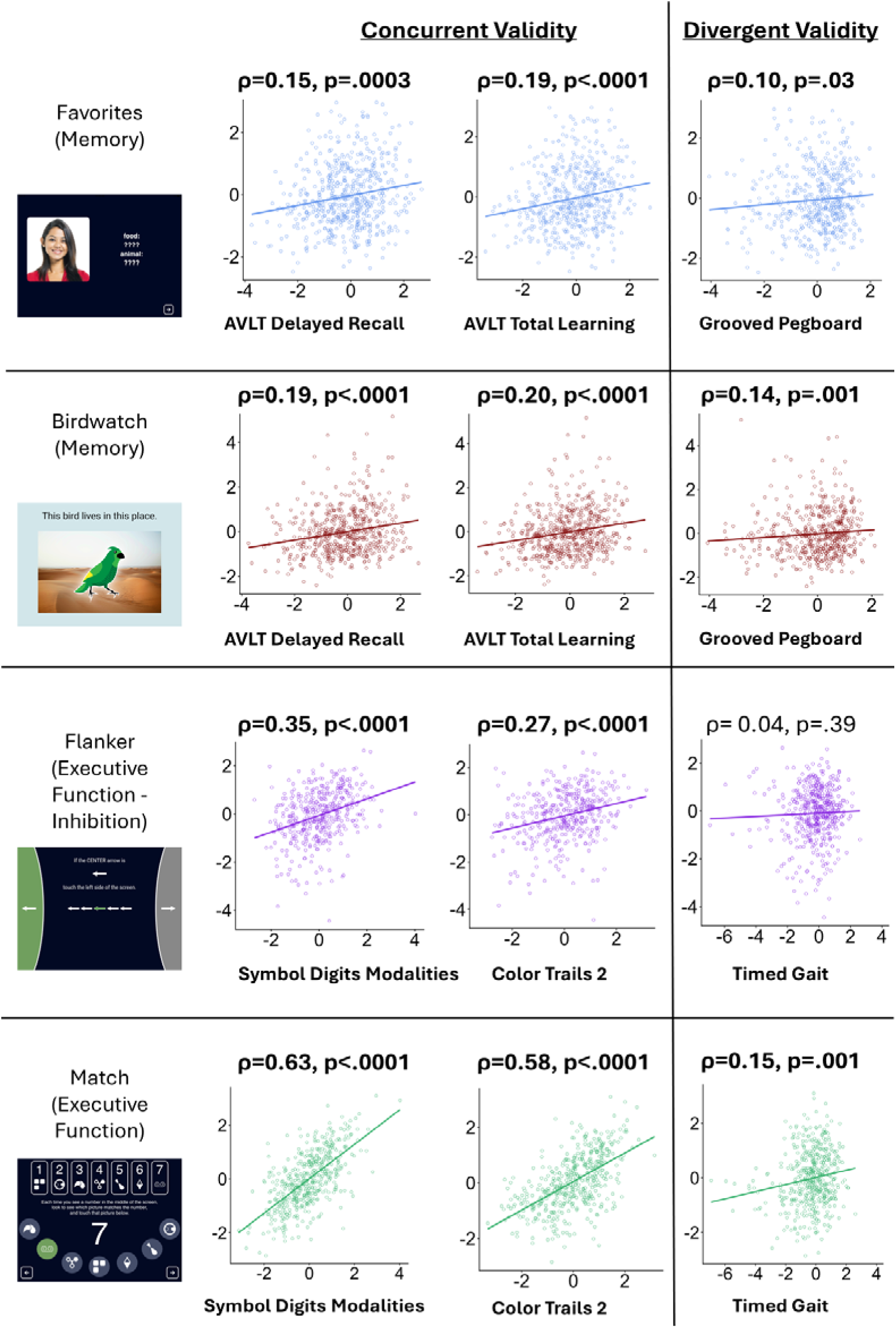
TabCAT Subtest Correlation with Reference-Standard Tests in Aligning and Non-aligning Domains. TabCAT tests demonstrated marginally stronger correlations with reference-standard tests in aligning cognitive domains than those in non-aligning domains. Abbreviations: AVLT = auditory verbal learning test

### Discrimination of Cognitive Impairment

We next evaluated the performance of the TabCAT Composite Score to discriminate objective cognitive impairment. The TabCAT Composite Score identified cognitively impaired from unimpaired individuals with an AUC of 0.77 (**Figure 4**). TabCAT demonstrated better discrimination of cognitive impairment among PLWH than among adults not living with HIV (**Figure 4**, PLWH: AUC 0.82 (95% CI 0.76-0.87); adults not living with HIV: AUC 0.71 (95% CI 0.63-0.78); DeLong test p=0.02).

**Figure 4.**
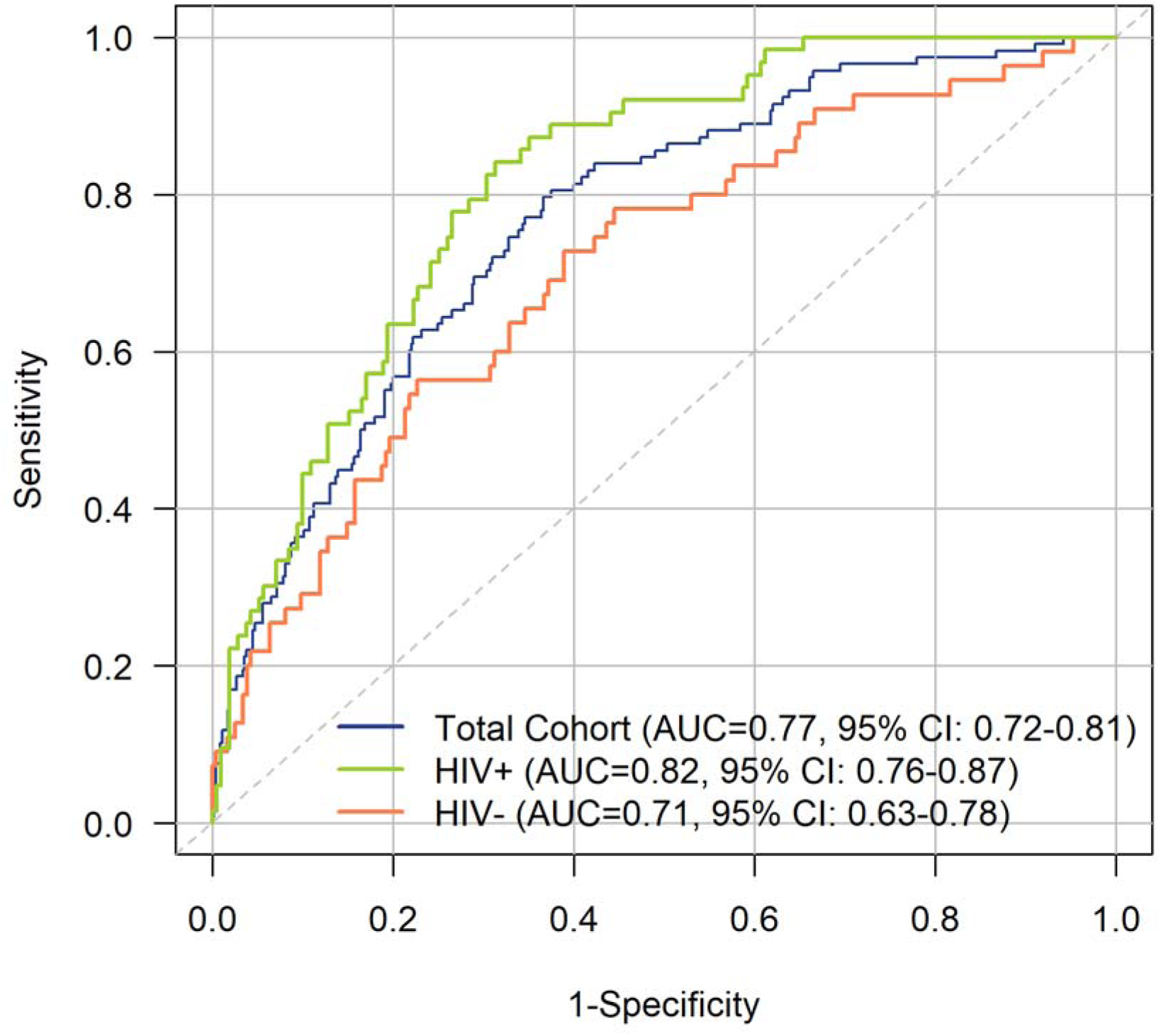
TabCAT Composite Score Performance in Discriminating Objective Cognitive Impairment. The TabCAT Composite Score in the total cohort (blue), then stratified by people living with HIV (green) and older adults not living with HIV (orange).

In sensitivity analyses, we assessed the discriminative performance of the TabCAT Composite Score using alternative criteria to define objective cognitive impairment. TabCAT performance was comparable across both thresholds (-1 and -1.5 SD) with Jak/Bondi criteria (**Supplementary Figure 4**). TabCAT had greater discriminative performance when using cut points of increasing severity in the Global Cognitive Composite Score to define cognitive impairment.

TabCAT Composite Score performance did not differ significantly according to the number of available TabCAT test scores used to derive the score. The AUC was 0.76 when all four tests were available (95% CI: 0.70–0.81), 0.80 when three tests were available (95% CI: 0.71–0.88), and 0.69 when two tests were available (95% CI: 0.43–0.95). Pairwise comparisons using DeLong tests showed no statistically significant differences between groups (2 vs. 3 tests: p=0.46; 2 vs. 4 tests: p=0.63; 3 vs. 4 tests: p=0.45).

Among individual TabCAT tests, Match had the strongest performance in distinguishing individuals with cognitive impairment (AUC 0.78) while Favorites, Birdwatch, and Flanker had comparable performance (AUC 0.63-0.67) (**Figure 5**). In post-hoc analyses, results were similar when limited only to participants who were literate (Match AUC 0.78, Favorites AUC 0.65, Birdwatch AUC 0.67, and Flanker AUC 0.62).

**Figure 5.**
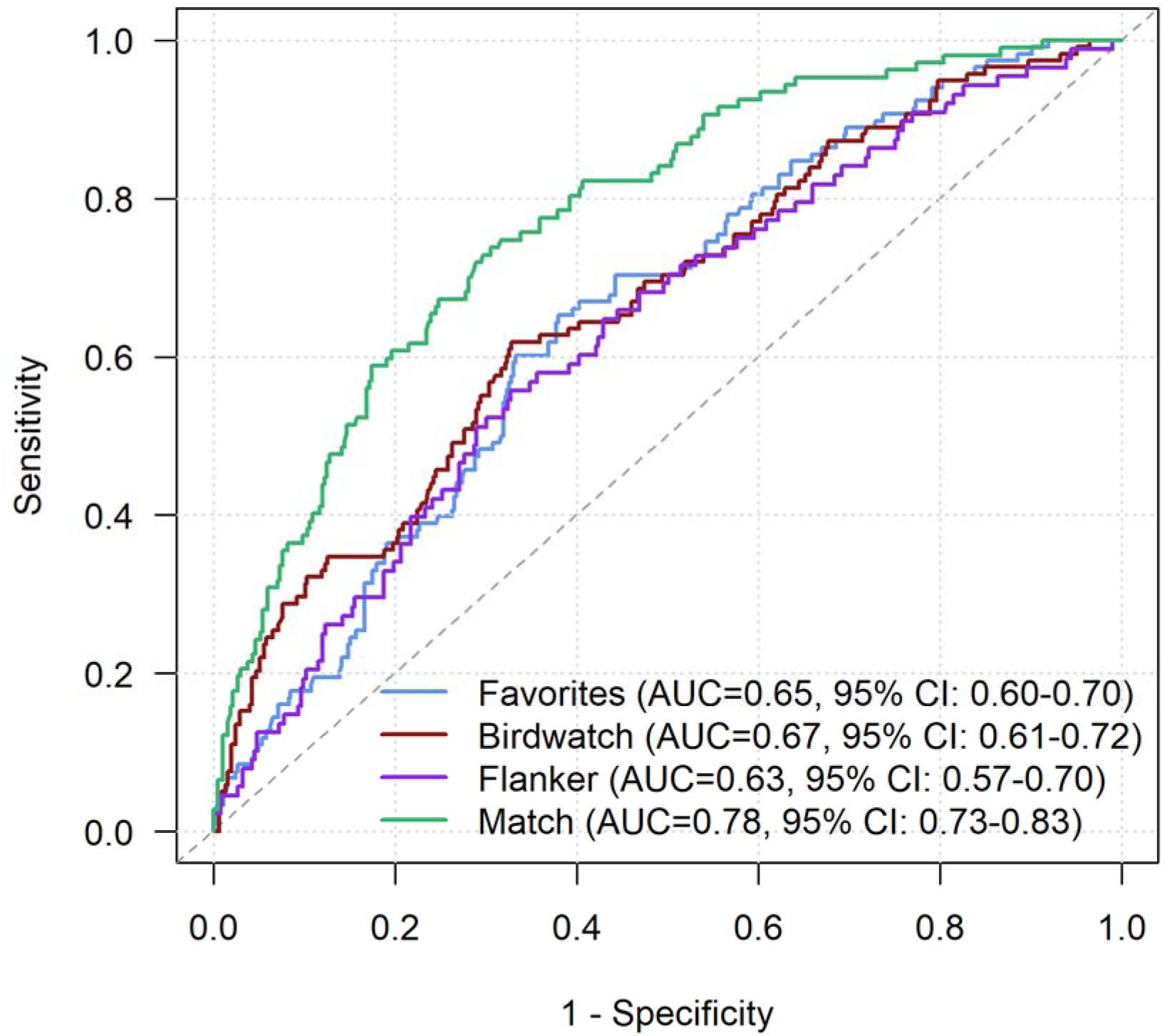
TabCAT Subtest Performance in Discriminating Objective Cognitive Impairment. TabCAT Match has the highest AUC in discriminating objective cognitive impairment, while TabCAT Favorites, Birdwatch, and Flanker perform comparably to each other.

## Discussion

There is a rapidly growing need for accessible, scalable, and validated brief cognitive assessment tools to identify cognitive impairment in older adults and older PLWH in sSA. In this study, we sought to translate, culturally adapt, and validate a novel brief tablet-based cognitive assessment tool compared to reference-standard pen-and-paper tests in a representative cohort of 563 older adults in southwestern Uganda, including well-treated PLWH. We found that the TabCAT battery demonstrated face, content, construct, concurrent, and criterion validity in measuring cognition and detecting objective cognitive impairment relative to the reference-standard testing. While additional studies in this cohort and others in sSA are needed to further validate the discriminative performance and generalizability of TabCAT, these results support its potential as a low-cost, brief cognitive assessment tool for detecting cognitive impairment in Uganda, including among older PLWH, and in populations with lower levels of education and literacy.

Our results are consistent with prior studies of TabCAT measures in other cohorts in the Global South, further supporting use in culturally diverse populations and across education levels. In a prior cross-sectional study in 146 participants in Cuba, TabCAT tests (Favorites, Match, and Line Orientation) outperformed the MoCA in discriminating between cognitively impaired and unimpaired adults (TabCAT AUC 0.95; MoCA AUC 0.83).^18^ A prior study in Nigeria evaluated the feasibility and demographic determinants of TabCAT tests (Favorites, Match, Line Length, and Animal Fluency) in 207 adults.^46^ TabCAT was shown to be feasible in both rural and urban populations in primary healthcare clinics with lower age, higher education, and rurality being associated with performance. Our study extends these findings by validating the TabCAT discriminative performance in a representative cohort in Uganda that includes older PLWH and participants unable to read. Additionally, our study included two TabCAT tests not previously validated in sSA: Flanker (executive function, inhibition) and Birdwatch (associative memory). Indeed, the newer Birdwatch test had similar-to-improved performance across measures relative to Favorites (associative memory), greater reported cultural appropriateness per focus groups, and improved ease of use. As a result, it has now been adopted as the preferred TabCAT memory test in subsequent UADCS annual study visits and continues to be collected with Flanker and Match.

Correlations between TabCAT tests and pen-and-paper tests in aligning domains were weaker than anticipated, particularly for memory. This may reflect differences in the constructs assessed: TabCAT tests (Birdwatch, Favorites) emphasize visually mediated associative memory, whereas AVLT measures verbal memory. Importantly, correlations were still higher with aligning than with non-aligning domains, supporting construct validity.

To develop regression-based normative scores, we evaluated demographic and clinical variables potentially associated with cognitive performance in cognitively unimpaired individuals. Age, sex, education level, and urbanicity were meaningfully associated with cognitive performance, similar to prior studies in sSA.^15,46,47^ In this cohort, literacy status (defined as ability to read/count) was associated with cognitive performance in multiple reference-standard and TabCAT tests, independent of education and other covariates. Prior studies in the Global North have demonstrated that little to no literacy is associated with worse cognitive test performance and increased risk of dementia, though not with more rapid cognitive decline.^48–50^ Data in the Global South and sSA, where a greater proportion of older individuals are unable to read,^51^ are notably lacking. A prior study in rural Tanzania found that illiteracy was associated with higher odds of scoring as “probable dementia” on screening tests, but not after clinical evaluation.^47^ Future studies should evaluate whether literacy is a potentially modifiable dementia risk factor or a confounding factor on cognitive testing in sSA.

Strengths of this study include rigorous translation and cultural adaptation of test materials prior to administering cognitive testing. Additionally, validity testing was performed in a large, representative cohort in rural Uganda. Testing was conducted by research staff (non-specialists) to evaluate whether TabCAT tests are scalable and applicable for use by non-specialists.

There are several limitations. First, although the cohort is among one of the oldest cohorts of PLWH in sSA, the mean age of 60 is still relatively young, and there may be less cognitive impairment present than predicted by Jak/Bondi criteria. Second, the reference-standard test battery was designed to assess HIV-associated cognitive impairment and related cognitive domains, rather than AD and other neurodegenerative causes of dementia. This may explain the higher performance among PLWH. To assess the implications of this potential bias, future studies will use a broader cognitive testing battery designed to evaluate ADRDs (including additional visuospatial and language tests) and will include clinical diagnoses of mild cognitive impairment and dementia (determined by subsequent multidisciplinary consensus diagnostic conferences) as the primary outcome. These assessments were added to UADCS study visits starting in 2025. Third, there were no regional cognitive datasets that could be used as a reference for normal cognitive functioning, and thus Z-scores were derived from cognitively unimpaired adults within the cohort. Fourth, cognitive test scores from the same UADCS study visits were used for both the reference-standard (2^nd^ year administered) and TabCAT tests (1^st^ year administered). While this was to enable collection and comparison of testing on the same day to minimize biases, there may be confounding from practice effects from reference-standard testing. Lastly, certain cognitive tests had a restricted sample size (e.g., TabCAT Flanker did not include participants who did not pass training trials), which could limit generalizability. Further validation in external cohorts is necessary.

In conclusion, this study supports a tablet-based cognitive assessment as a promising brief digital cognitive assessment tool to measure cognitive performance and detect cognitive impairment in older aged adults in Uganda, including in PLWH and in a population-representative sample with varying levels of education and literacy. After translation and cultural adaptation, TabCAT measures were regarded to have acceptable face and content validity, and demonstrated construct, concurrent, and criterion validity in measuring cognition. The TabCAT Composite Score demonstrated good discriminative ability in identifying objective cognitive impairment relative to reference-standard testing. Futures studies should assess TabCAT and other digital assessment tools in other diverse, international cohorts and evaluate performance in detecting longitudinal cognitive decline and dementia.

## Data Availability

All data produced in the present study are available upon reasonable request to the Corresponding Author.

## Acknowledgements

We would like to thank the research participants in Uganda and the study team from Mbarara and Kabwohe in Uganda for making this work possible.

## Funding

This work was supported by the US National Institutes of Health National Institute of Aging (NIH NIA; R56AG087790, R01AG059504, K08AG090815, P30AG066546) and a research grant from the Alzheimer’s Association and The Michael J. Fox Foundation for Parkinson’s Research (26BFDN-1579177A). ACT acknowledges additional support from NIH K24DA061696. The contents of this manuscript are solely the responsibility of the authors and do not necessarily represent the official views of the NIH.

## Authorship

R.V. contributed to study design, data analyses, original writing, and review and editing. G.H. and C.W. contributed to study design, data analyses, and writing review and editing. R.P., N.N., Z.R., F. A., E. T., M.G., E. P., C. R., S. S. H., A. T., J. S., A. W., S. S., S. O., and S. A. contributed to conceptualization, study design, study implementation, and writing review and editing. D. S., K. L. P., E. T. and M. J. S. contributed to conceptualization, study design, study implementation, data analyses, and writing review and editing. E.V. contributed to review and editing. J. A. T. contributed to conceptualization, study design, study implementation, data analyses, original writing, and review and editing.

## Competing Interest

ACT reports receiving financial honoraria from Elsevier (for his work as Co-Editor in Chief of *SSM – Mental Health*) and BMJ Publishing Group Ltd. (for his work as Clinical Editorial Advisor of *The BMJ*). The other authors have no relevant financial or non-financial conflicts of interest to disclose.

---

**Supplementary Table 1.**
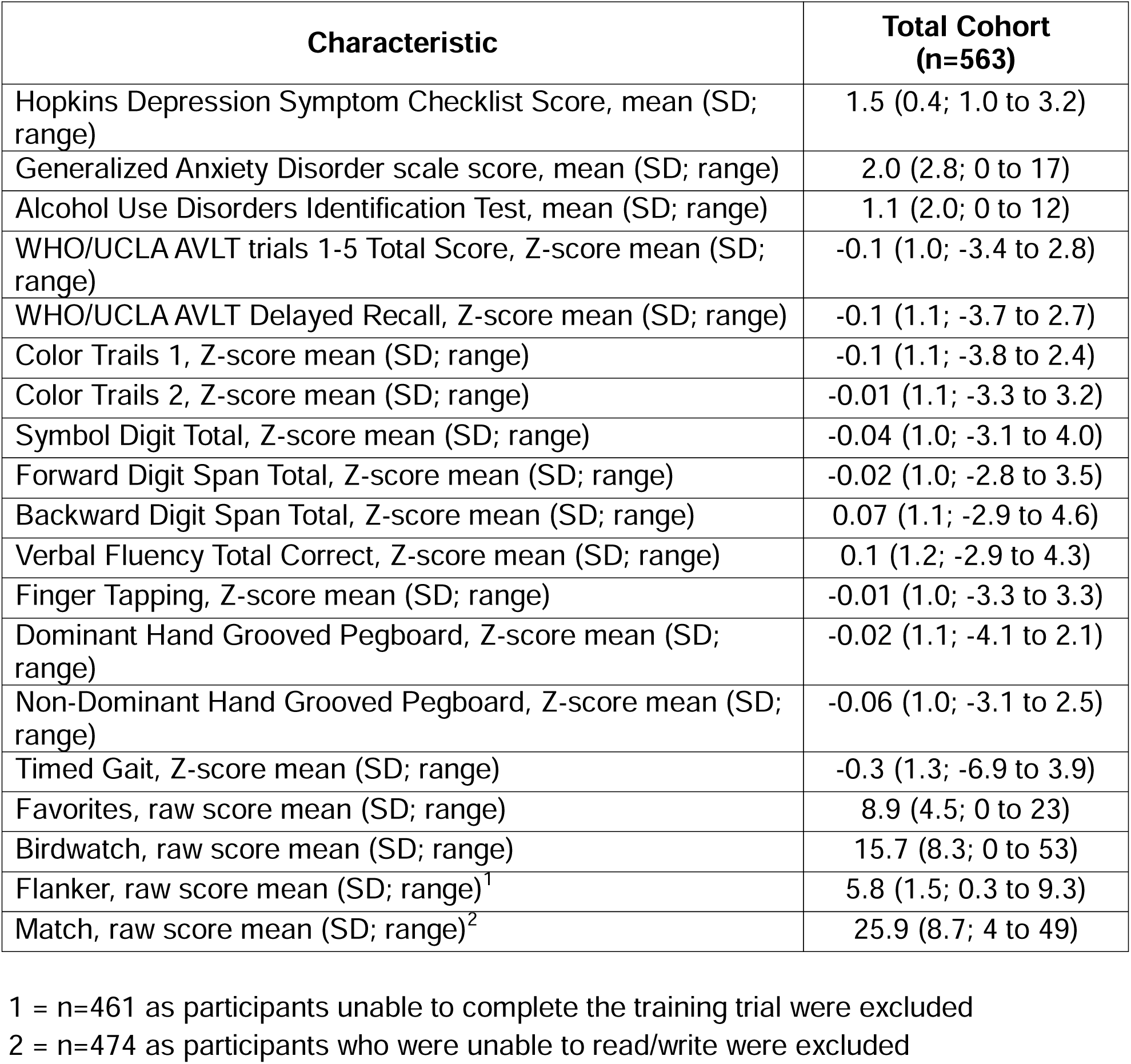
Participant Characteristics in the Uganda Aging and Dementia Cohort Study.

| **Characteristic** | **Total Cohort**<br>**(n=563)** |
| --- | --- |
| Hopkins Depression Symptom Checklist Score, mean (SD; range) | 1.5 (0.4; 1.0 to 3.2) |
| Generalized Anxiety Disorder scale score, mean (SD; range) | 2.0 (2.8; 0 to 17) |
| Alcohol Use Disorders Identification Test, mean (SD; range) | 1.1 (2.0; 0 to 12) |
| WHO/UCLA AVLT trials 1-5 Total Score, Z-score mean (SD; range) | -0.1 (1.0; -3.4 to 2.8) |
| WHO/UCLA AVLT Delayed Recall, Z-score mean (SD; range) | -0.1 (1.1; -3.7 to 2.7) |
| Color Trails 1, Z-score mean (SD; range) | -0.1 (1.1; -3.8 to 2.4) |
| Color Trails 2, Z-score mean (SD; range) | -0.01 (1.1; -3.3 to 3.2) |
| Symbol Digit Total, Z-score mean (SD; range) | -0.04 (1.0; -3.1 to 4.0) |
| Forward Digit Span Total, Z-score mean (SD; range) | -0.02 (1.0; -2.8 to 3.5) |
| Backward Digit Span Total, Z-score mean (SD; range) | 0.07 (1.1; -2.9 to 4.6) |
| Verbal Fluency Total Correct, Z-score mean (SD; range) | 0.1 (1.2; -2.9 to 4.3) |
| Finger Tapping, Z-score mean (SD; range) | -0.01 (1.0; -3.3 to 3.3) |
| Dominant Hand Grooved Pegboard, Z-score mean (SD; range) | -0.02 (1.1; -4.1 to 2.1) |
| Non-Dominant Hand Grooved Pegboard, Z-score mean (SD; range) | -0.06 (1.0; -3.1 to 2.5) |
| Timed Gait, Z-score mean (SD; range) | -0.3 (1.3; -6.9 to 3.9) |
| Favorites, raw score mean (SD; range) | 8.9 (4.5; 0 to 23) |
| Birdwatch, raw score mean (SD; range) | 15.7 (8.3; 0 to 53) |
| Flanker, raw score mean (SD; range)<sup>1</sup> | 5.8 (1.5; 0.3 to 9.3) |
| Match, raw score mean (SD; range)<sup>2</sup> | 25.9 (8.7; 4 to 49) |

1 = n=461 as participants unable to complete the training trial were excluded
2 = n=474 as participants who were unable to read/write were excluded

**Supplementary Table 2.** Participant Characteristics in the Normative Sample.

| **Characteristic** | **Total Cohort**<br>**(n=231)** |
| --- | --- |
| Age in years, mean (SD; range) | 59.8 (5.9; 50 to 84) |
| Female, n (%) | 116 (50%) |
| Education, n (%) |  |
| No schooling | 29 (13%) |
| Some primary school | 84 (36%) |
| Completed primary education | 54 (23%) |
| Some secondary school | 39 (17%) |
| Secondary school or higher | 25 (11%) |
| Literacy, n (%) |  |
| No | 30 (13%) |
| Yes | 201 (87%) |
| Urbanicity, n (%) |  |
| Semi-urban (Mbarara) | 93 (40%) |
| Rural (Kabwohe) | 138 (60%) |
| HIV Positive, n (%) | 0 (0%) |
| Global Cognitive Composite, Z-score mean (SD; range) | -0.002 (0.6; -1.7 to 1.9) |
| TabCAT Cognitive Composite, Z-score mean (SD; range) | -0.01 (0.7; -1.8 to 2.1) |
| Favorites, Z-score mean (SD; range) | -0.001 (1.0; -1.9 to 2.9) |
| Birdwatch, Z-score mean (SD; range) | -0.001 (1.0; -2.4 to 3.3) |
| Flanker, Z-score mean (SD; range)<sup>1</sup> | 0.003 (1.0; -3.3 to 2.6) |
| Match, Z-score mean (SD; range)<sup>2</sup> | -0.01 (1.0; -2.9 to 2.9) |

1 = n=194 as participants unable to complete the training trial were excluded
2 = n=201 as participants who were unable to read/write were excluded

**Supplementary Table 3.**
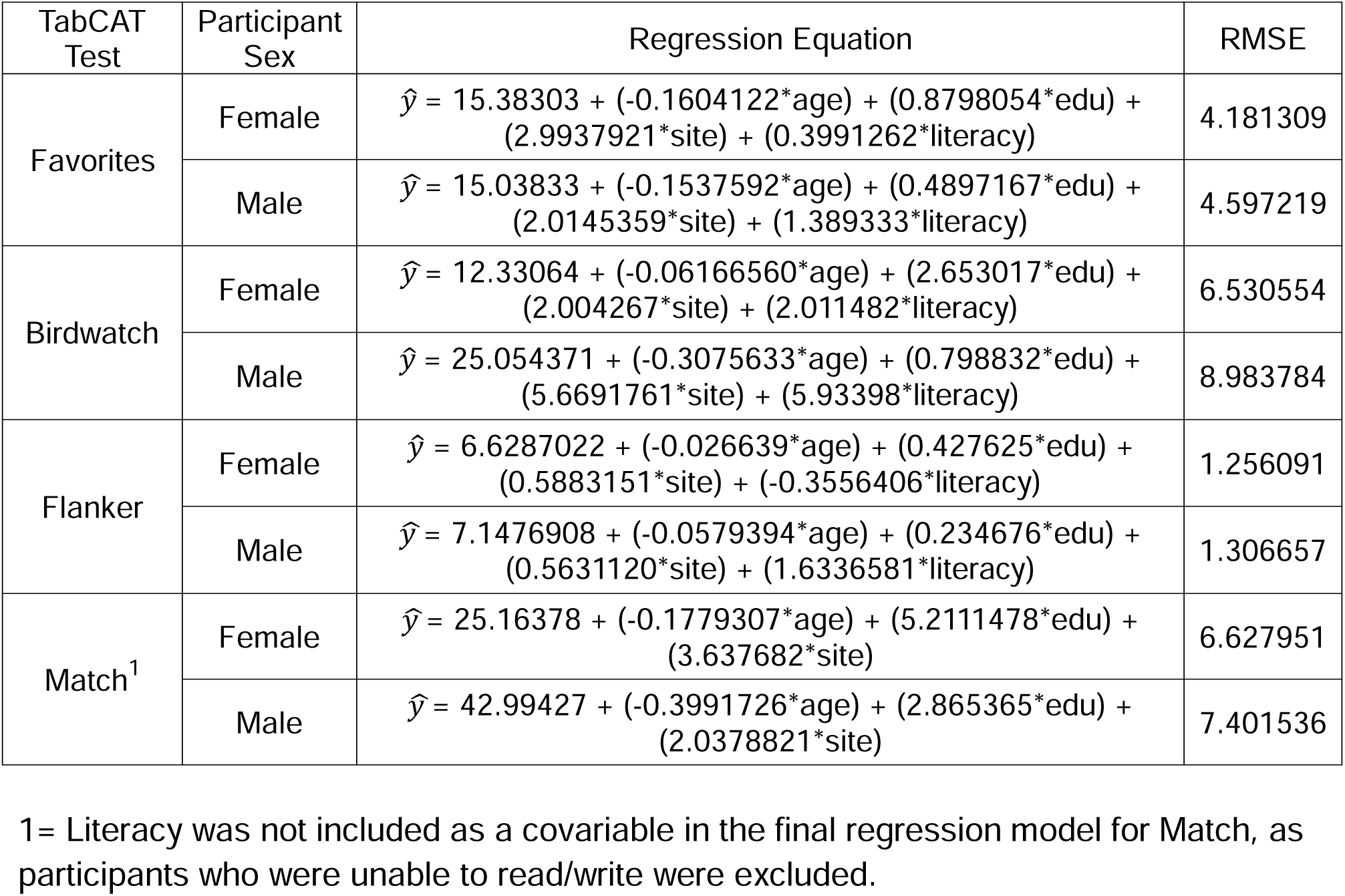
Regression Models for Calculating Z-Scores for TabCAT Tests.

**Supplementary Table 4.** Floor and Ceiling Effects Among TabCAT Tests in the UADCS by Literacy Status.

| TabCAT Test | Literate<br>N=474 |  |  |  |  | Not Literate<br>N=89 |  |  |  |  |
| --- | --- | --- | --- | --- | --- | --- | --- | --- | --- | --- |
|  | # at Min,<br>N (%) | Q1 | Median<br>(SD) | Q3 | # at Max,<br>N (%) | # at Min,<br>N (%) | Q1 | Median<br>(SD) | Q3 | # at Max,<br>N (%) |
| Favorites | 3 (0.6%) | 6 | 9 (4.6) | 12 | 0 (0%) | 0 (0%) | 5 | 7 (4.2) | 10 | 0 (0%) |
| Birdwatch | 0 (0%) | 11 | 15 (8.5) | 21 | 0 (0%) | 1 (1.1%) | 8 | 10 (4.7) | 13 | 0 (0%) |
| Flanker<sup>1</sup> | 0 (0%) | 5 | 6.1<br>(1.4) | 6.9 | 0 (0%) | 0 (0%) | 3.2 | 5 (1.7) | 6 | 0 (0%) |
| Match<sup>2, 3</sup> | 0 (0%) | 20 | 26 (8.7) | 32 | - | - | - | - | - | - |

1= Participants who were unable to pass the Flanker trial were excluded from this analysis (n=414 among literate participants; n=47 among non-literate participants).
2= Match scores are continuous and do not have a prespecified maximum score.

**Supplementary Figure 1.**
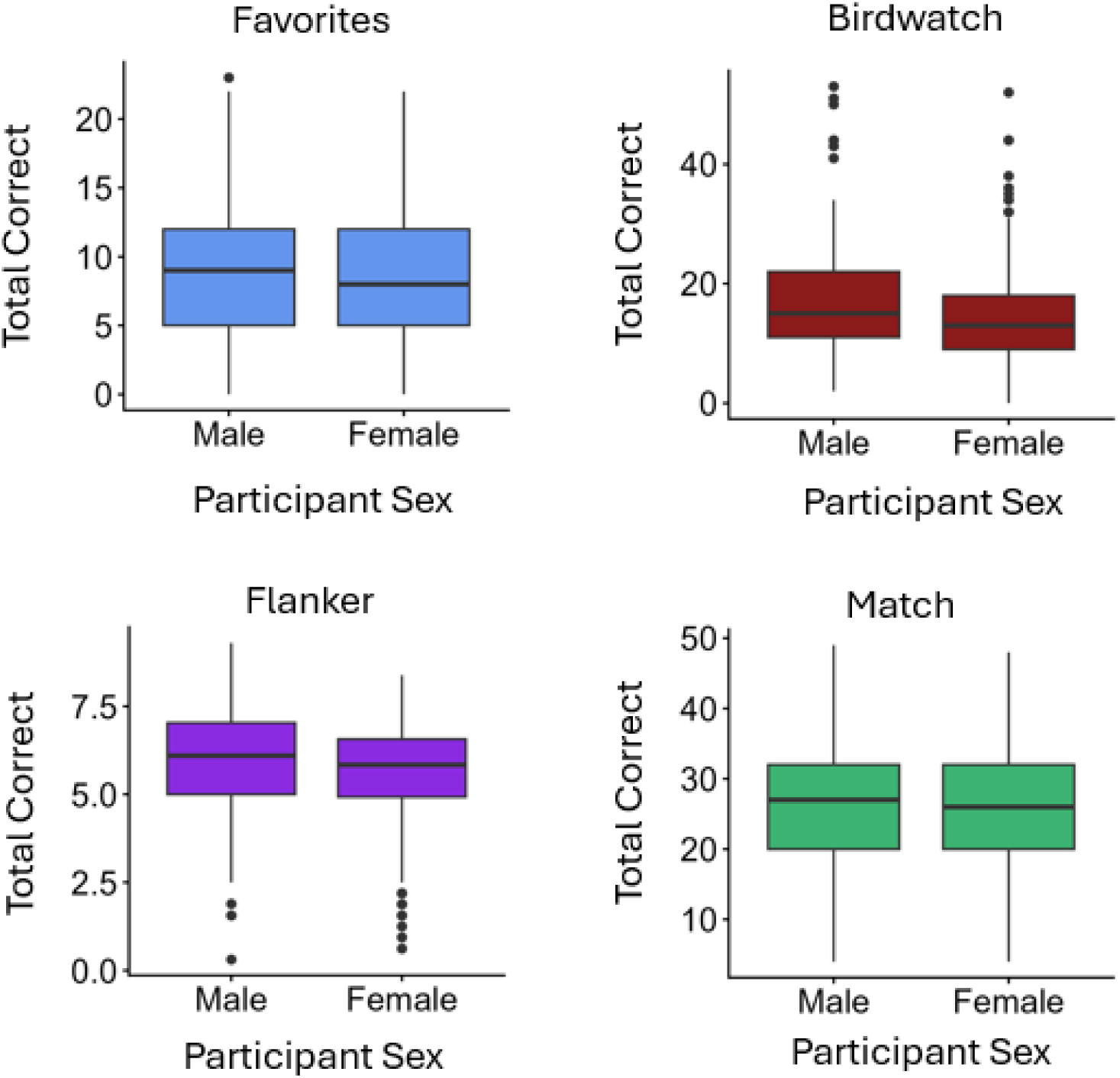
TabCAT test performance by participant sex. Male participants generally exhibited higher median scores on TabCAT tests.

**Supplementary Figure 2.**
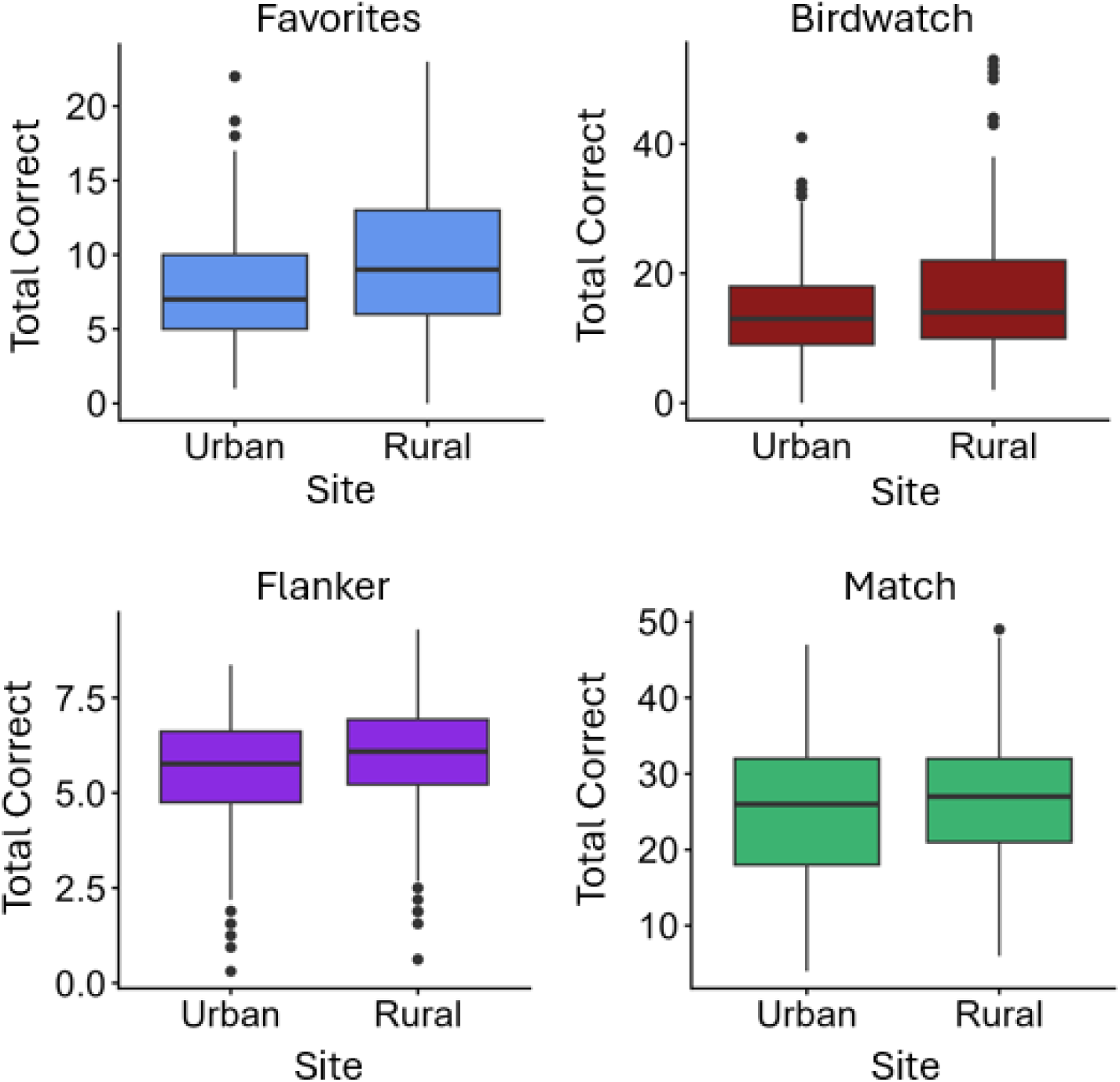
TabCAT test performance by urbanicity. Participants at the rural Kabwohe site generally exhibited higher performance on TabCAT tests compared to those at the semi-urban Mbarara site.

**Supplementary Figure 3.**
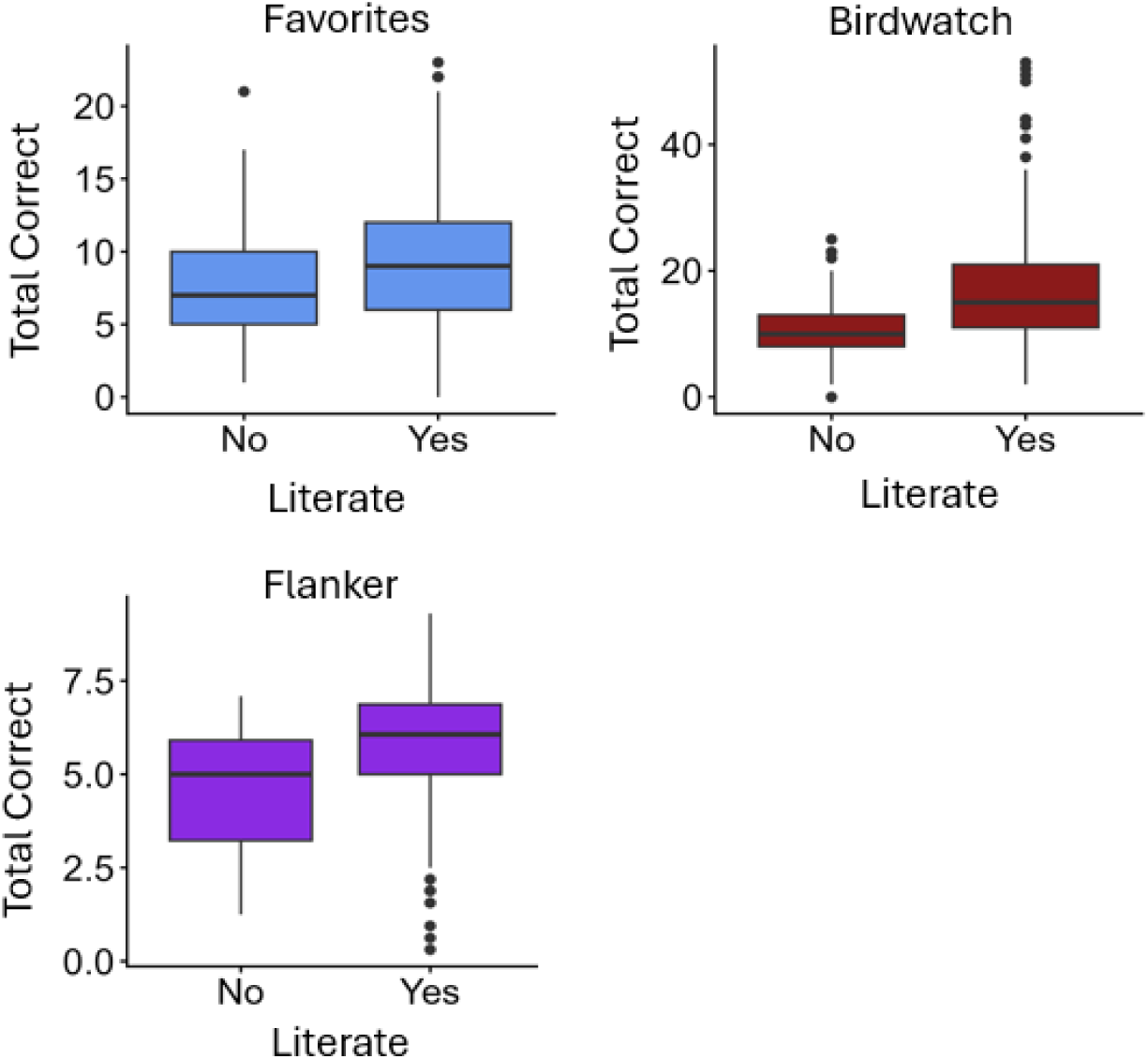
TabCAT test performance by literacy. Participants who were able to read and write generally exhibited higher performance on TabCAT tests compared participants who were unable to read and write. Match results were omitted as participants who were unable to read/write were excluded.

**Supplementary Figure 4.**
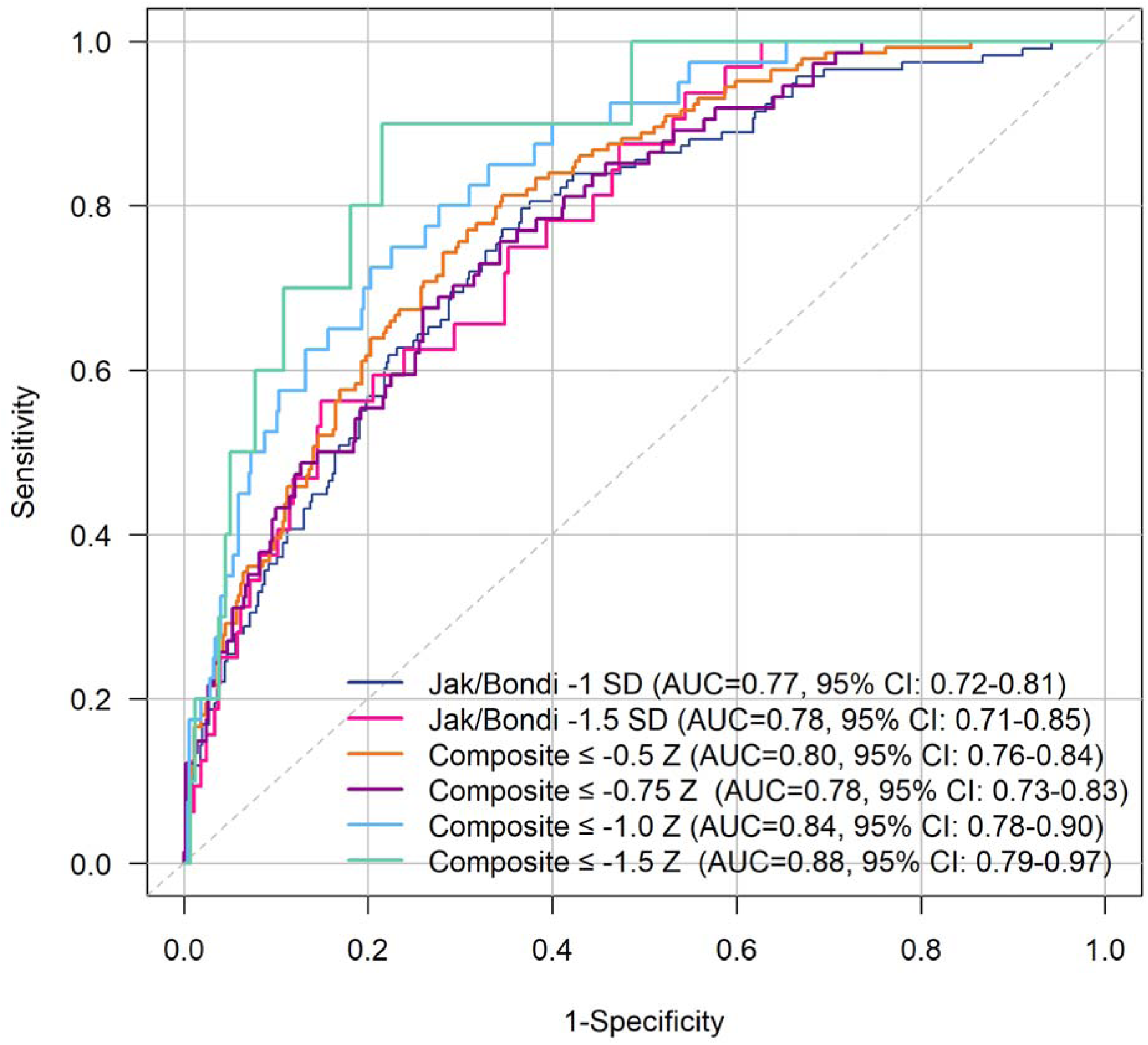
TabCAT Composite Performance in Discriminating Objective Cognitive Impairment Defined with Differing Criteria. TabCAT discriminative performance increases with more stringent thresholds for cognitive impairment.

